# Leveraging Large Language Models in Gynecologic Oncology: A Systematic Review of Current Applications and Challenges

**DOI:** 10.1101/2024.08.08.24311699

**Authors:** Aya Mudrik, Abraham Tsur, Girish N Nadkarni, Orly Efros, Benjamin S Glicksberg, Shelly Soffer, Eyal Klang

## Abstract

**Rationale and Objectives:** Over the past year, studies have been conducted to evaluate the performance of Large Language Models (LLMs), such as ChatGPT, in the fields of gynecologic oncology. This review aims to analyze the applications and risks associated with using LLMs in this specialized field.

**Materials and Methods:** This systematic review was performed in adherence to the Preferred Reporting Items for Systematic Reviews and Meta-Analyses (PRISMA) guidelines, incorporating elements from the diagnostic test accuracy extension and the CHARMS checklist for reviews of prediction models. A systematic literature search was executed on July 17, 2024, across PubMed, Web of Science, and Scopus databases. We focused on identifying original research that integrates LLMs with gynecologic oncology. We assessed the risk of bias using the adapted QUADAS-2 criteria.

**Results:** Our search identified eight studies that met our criteria, focusing on healthcare education, clinical practice, and medical code generation. These studies revealed variability in ChatGPT’s performance across different applications. It excelled in genetic testing and counseling, achieving 97% accuracy rate. However, its performance in cervical cancer prevention was less robust, with an accuracy of 83%. While one study demonstrated ChatGPT’s high adherence to quality guidelines, another noted that established guidelines significantly outperformed ChatGPT’s outputs. Additionally, code generation using tools like Google Bard and RoBERTa have shown potential to improve accuracy in clinical predictions and quality assurance. For example, Natural Language Processing (NLP) assisted by RoBERTa (based on Google’s BERT model) has improved the prediction of residual disease in women with advanced epithelial ovarian cancer following cytoreductive surgery. Despite these advancements, challenges related to consistency, specificity, and personalization persist, underscoring the necessity for continuous enhancement of these technologies.

**Conclusion:** LLMs demonstrate inconsistent performance in gynecologic oncology. These findings emphasize the need for continuous evaluation of these models before they are implemented clinically.

## INTRODUCTION

In recent years, LLMs such as OpenAI’s GPT, have demonstrated remarkable capabilities in understanding and generating human language, opening new avenues for their application in healthcare.^1^ Oncologic care, which encompasses early detection, precise staging, tailored therapeutic strategies, and ongoing patient support, can benefit by the data processing capabilities of LLMs.^2^ ^3^

Despite their promising applications, LLMs also present significant challenges within the medical field. These models require large training data, which raises concerns about patient privacy and data security.^4^ This issue is particularly critical in gynecologic oncology, a field that deals with sensitive and complex conditions. Furthermore, LLMs may lack the capability to account for the nuanced, individual patient contexts that are crucial in medical decision-making, potentially leading to oversimplified or inappropriate treatment recommendations. As LLMs gain prevalence in healthcare, the need to rigorously evaluate their applications grows.^5^

In a review published in May 2023 that explored the application of ChatGPT in obstetrics and gynecology, no studies were found that specifically evaluated ChatGPT’s effectiveness in gynecologic oncology.^6^ However, since that review, several articles have emerged in the fields of gynecology, obstetrics, and particularly gynecologic oncology. These recent publications show advancements and explore the potential of LLMs, especially in gynecologic oncology, demonstrating their diverse applications across medical education, clinical practice, and medical code generation.

We systematically reviewed the current research regarding the integration of LLMs in gynecologic oncology, focusing on possible clinical applications and limitations.

### Key Concepts and Terminology in Large Language Model

In **Figure 1**, we present a hierarchy diagram of artificial intelligence (AI) terms.

**Figure 1.**
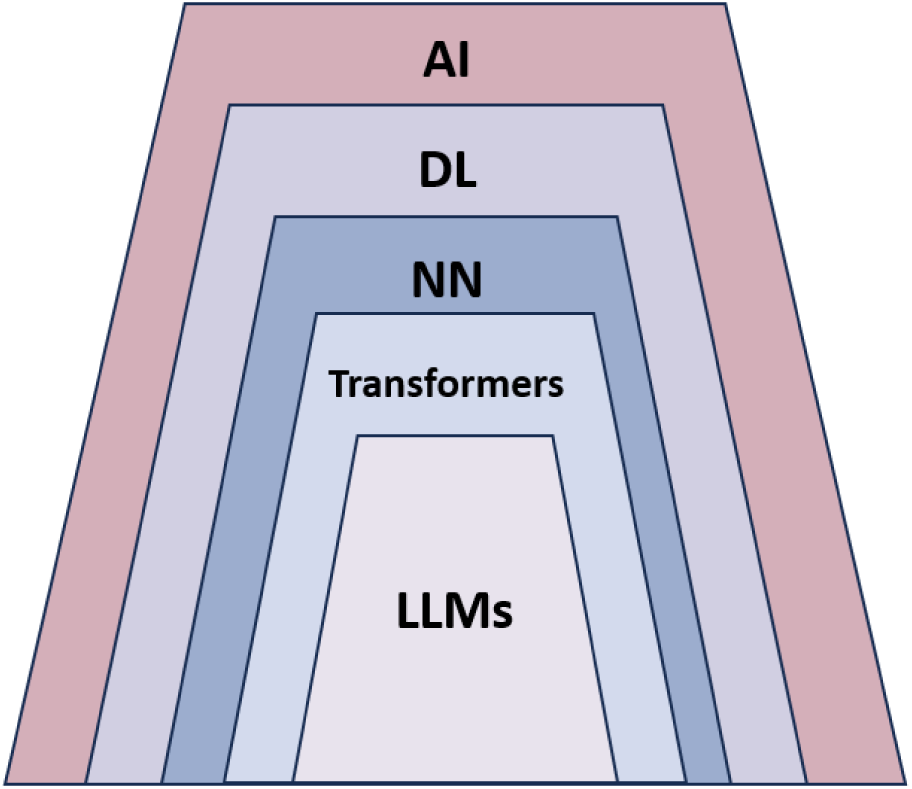
A hierarchy diagram of AI term

#### AI (Artificial Intelligence)

AI replicates human cognitive functions using machines, especially computer systems.^7^

#### Deep Learning (DL)

Deep Learning is a specialized branch of artificial intelligence (AI) that enables computers to process and interpret data using models called neural networks. These networks, inspired by the human brain’s structure, excel at identifying patterns across various data types such as images, text, and audio. This capability allows for generating insights and predictions.^7^

#### Neural Networks

A Neural Network is a deep learning system inspired by the biological neural networks. It consists of many small, repeating units called “neurons” or “nodes”. Each neuron is like a simple logistic regression unit, which takes in inputs, performs calculation, and produces an output. These neurons are connected to each other in layers, allowing the network to process and represent increasingly complex information. By combining multiple layers of neurons, the network can learn to recognize patterns, make predictions, and solve complex problems^8^.

#### Transformer Models

Transformer models are a type of advanced neural network designed to analyze sequential data, such as sentences, and understand context and meaning. They employ a mechanism called self-attention to examine relationships between elements in the data and assess how they interact and influence each other.^9^

For example, consider the sentence: “The patient’s cervical biopsy revealed high-grade squamous intraepithelial lesions”.

In this sentence, the self-attention mechanism would allow the model to understand that:

- *“The patient”* is the subject receiving the biopsy
- *“cervical biopsy”* is the procedure performed
- *“revealed”* indicates the result of the procedure
- *“high-grade squamous intraepithelial lesions”* is the diagnosis.

The self-attention mechanism helps the model to focus on the relationships between these elements, even though they are separated by other words in the sentence.

#### Large Language Models (LLMs)

These are complex systems usually composed of multiple layers of transformer models. LLMs are trained on vast amounts of data, enabling them to perform a range of tasks with high proficiency, including text recognition, translation, prediction, and generation. The extensive training allows LLMs to develop a deep understanding of language patterns and relationships. Notable LLMs include transformer-based models like GPT (Generative Pre-trained Transformer) by openAI, LLaMA (Large Language Model Meta AI) by Meta, Gemini by Google, and Claude by Anthropic, which have achieved state-of-the-art results in various NLP tasks.^10^ In **Figure 2**, we present a diagram of the way LLMs work.

**Figure 2.**
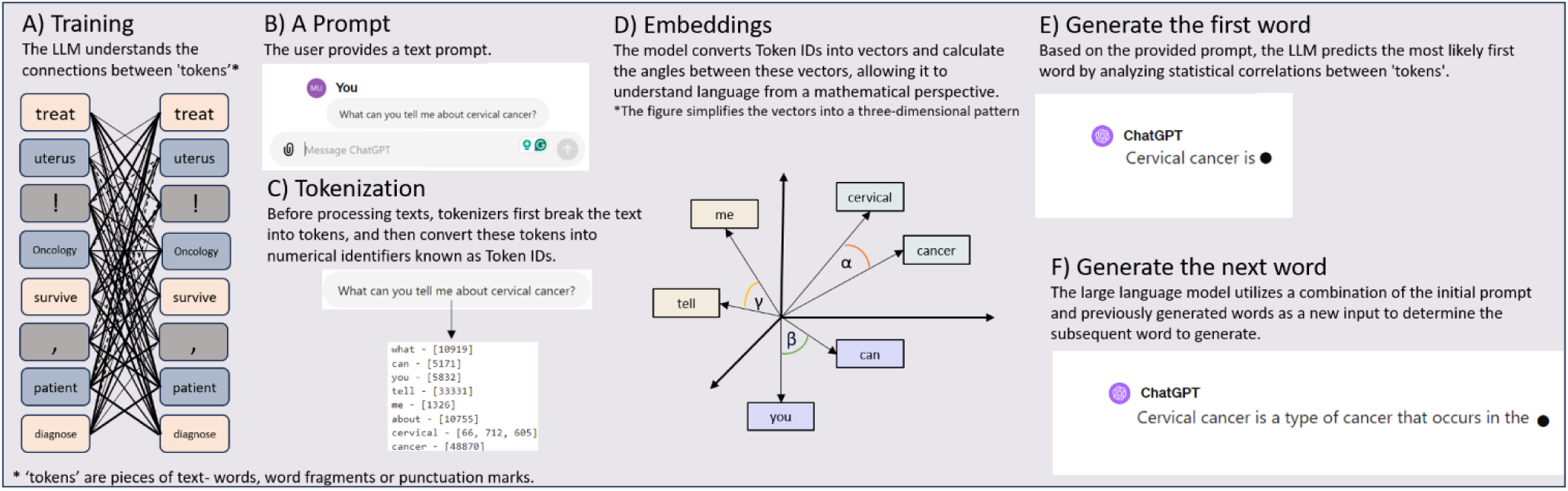
Diagram of the way LLMs work

## METHODS

### Search Strategy

This systematic review was performed in adherence to the Preferred Reporting Items for Systematic Reviews and Meta-Analyses (PRISMA) guidelines,^11^ incorporating elements from the diagnostic test accuracy extension^12^ and the CHARMS checklist^13^ for reviews of prediction models.

A systematic literature search was executed on July 17, 2024, across PubMed, Web of Science, and Scopus databases. We focused on identifying original research that integrates LLMs with gynecologic oncology, employing a set of specifically curated search terms detailed in the Supplementary Materials (“Detailed Search Strategies”).

The scope of our search was confined to peer-reviewed publications in English from December 1, 2022, onward, coinciding with the advent of ChatGPT and the broader release of LLMs. We excluded non-original studies, articles unrelated to the direct application of LLMs in gynecologic oncology, and conference abstracts. Additionally, references from selected articles were examined to capture any pertinent studies missed in the initial search.

The study is registered in the PROSPERO database (CRD42024555844).

### Study Selection

Initial screening of titles and abstracts was conducted independently by two reviewers (AM and SS), with eligibility based on predefined inclusion criteria. Any ambiguities were resolved through full-text assessments. Discrepancies during any stage of the selection process were resolved through consultation with a third reviewer (EK).

### Data Extraction

Data extraction was independently performed by the same two reviewers (AM and SS) using a standardized form designed for this review. Extracted data included publication year, types of LLMs utilized, study objectives, sample sizes, primary outcomes, and noted limitations.

### Quality Assessment and Risk of Bias

The risk of bias within the evaluated studies was assessed using an adapted version of the Quality Assessment of Diagnostic Accuracy Studies (QUADAS-2) criteria.^14^

### Data Synthesis

Given the heterogeneity in study designs and outcomes, we opted for a narrative synthesis over a meta-analysis. This approach allowed us to cohesively summarize the diverse applications, benefits, and challenges associated with the use of LLMs in gynecologic oncology, as reported by the included studies.

## RESULTS

A total of 135 articles were retrieved in the initial search. After exclusion **(Supplementary** Figure 1), 8 studies evaluating the application of LLMs in gynecology oncology were included.

The characteristics of the studies are presented in **Table 1**. Objectives, reference standards, sample sizes, and main findings are presented in **Table 2**. The included studies spanned multiple categories, including medical education, clinical practice, and medical code generation (**Figure 3**). The studies varied in their objectives and methodologies, covering a range of topics within gynecologic oncology. Topics included ovarian cancer, cervical cancer, endometrial cancer, postoperative instructions, palliative care scenarios, quality assurance audits, genetic testing, and the prediction of residual disease.

**Figure 3.**
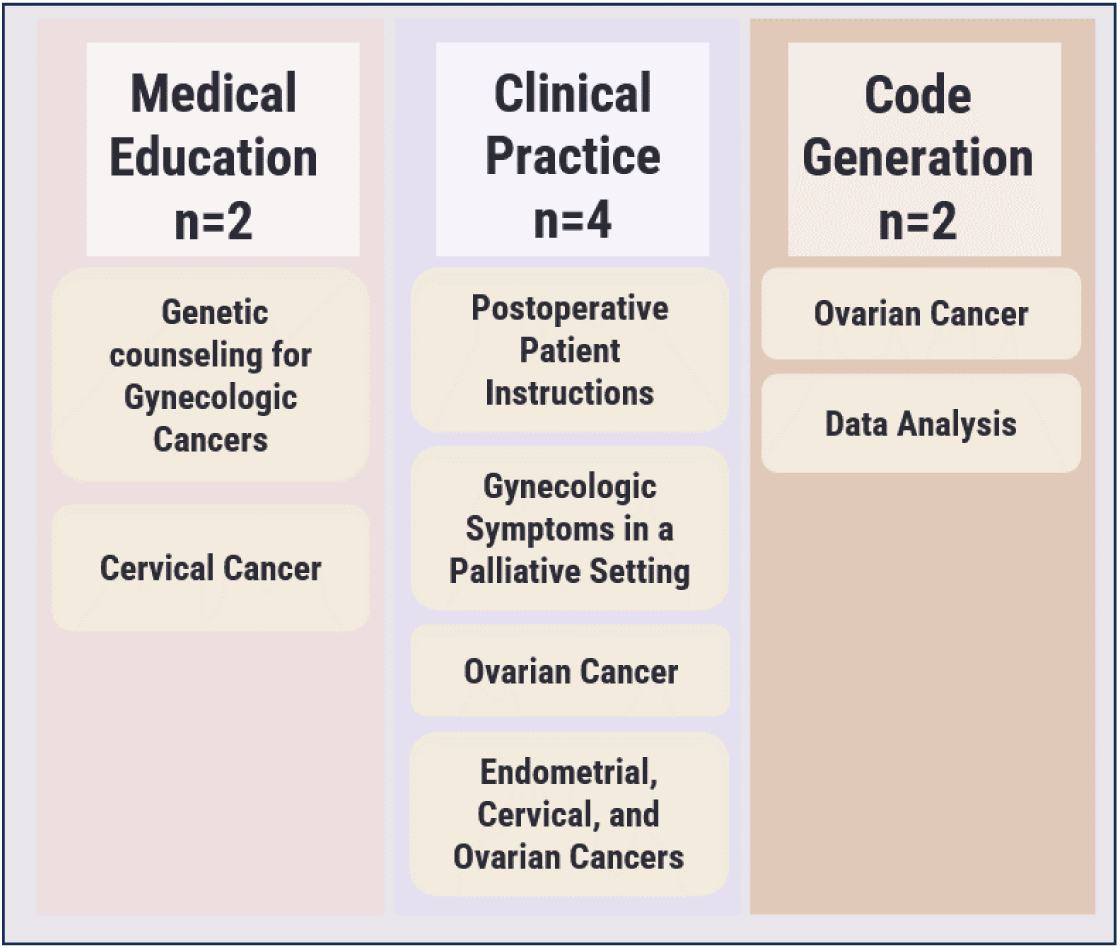
Applications of LLMs in Gynecologic Oncology in the Articles Reviewed

**Table 1.** Details about the reviewed articles.

| **Group** | **Title** | **First Author** | **Journal/Book** | **Year** |
| --- | --- | --- | --- | --- |
| **clinical practice** | Will I soon be out of my job? Quality and guideline conformity of ChatGPT therapy suggestions to patient inquiries with gynecologic symptoms in a palliative setting | Braun EM. et al. | Archives of Gynecology and Obstetrics | **2024** |
|  | The Consistency and Quality of ChatGPT Responses Compared to Clinical Guidelines for Ovarian Cancer: A Delphi Approach | Dario Piazza. et al. | Current Oncology | **2024** |
|  | Evaluation of ChatGPT's Potential in Tailoring Gynecological Cancer Therapies | Krückel A. et al. | In Vivo | **2024** |
|  | ChatGPT compared with Google Search and healthcare institution as sources of postoperative patient instructions after gynecological surgery | Meyer R. et al. | British journal of obstetrics and gynaecology (BJOG) | **2024** |
| **code generation** | RoBERTa-Assisted Outcome Prediction in Ovarian Cancer Cytoreductive Surgery Using Operative Notes | Laios A. et al. | Cancer Control | **2023** |
|  | Can natural language processing be effectively applied for audit data analysis in gynaecological oncology at a UK cancer centre? | McGowan M. et al. | International Journal of Medical Informatics | **2024** |
| **medical education** | Let's chat about cervical cancer: Assessing the accuracy of ChatGPT responses to cervical cancer questions | Hermann CE. et al. | Gynecologic Oncology | **2023** |
|  | ChatGPT accurately performs genetic counseling for gynecologic cancers | Patel JM. et al. | Gynecologic Oncology | **2024** |

**Table 2.** Methods and results of the reviewed articles.

| Group | First Author | LLM Type | objective | Reference standard | sample size | main findings |
| --- | --- | --- | --- | --- | --- | --- |
| clinical practice | Braun EM. et al. | GPT-3.5 | Evaluate the recommendations of GPT-3.5 about the possible therapy of gynecological leading symptoms in a palliative situation. | Five experts in palliative care and gynecologic oncologist and common guidelines | 10 queries | Overall rate of GPT-3.5 responses- 4.1/5 |
| clinical practice | Dario Piazza. et al. | GPT-3.5<br>GPT-4.0 | Investigate the consistency and quality of responses that ChatGPT generates regarding clinical queries about ovarian cancer | Italian Association of Medical Oncology (AIOM) guidelines for ovarian cancer and an expert panel of healthcare professionals | eight clinical questions | AIOM guidelines significantly outscored the GPT models, with no significant differences among the different GPT models. |
| clinical practice | Krückel A. et al. | GPT-3.5 | Evaluates ChatGPT's ability to provide therapy recommendations for gynecological malignancies | Answer Scoring System modified according to Lukac. et al. | Data collected during the routine clinical care of 11 patients. | (Scores ranged from a minimum of -1 point to a maximum of +2 points)<br>Ovarian Cancer- 0.75<br>Cervical Cancer- 0.7<br>Endometrial Cancer- 1.5 |
| clinical practice | Meyer R. et al. | GPT-3.5 | Study the value of ChatGPT generated postoperative instructions for gynecological procedures. | Google Search and authentic hospital discharge instructions for the surgical procedures and 2 researchers | 5 common gynecological procedures | Understandability<br>ChatGPT- 92%<br>Google- 83%<br>Institution- 87.5% (non-significant)<br>Actionability:<br>ChatGPT- 60%<br>Google- 70%<br>Institution- 80% (GPT vs Institution is significant)<br>Total:<br>ChatGPT- 82%<br>Google- 82%<br>Institution- 85.5% (non-significant) |
| code generation | Laios A. et al. | RoBERTa | Determine whether natural language processing (NLP) of unstructured operative notes improves the prediction of residual disease in women with advanced epithelial ovarian cancer following cytoreductive surgery. | Traditional clinical prediction models | 555 cases of epithelial ovarian cancer | Accuracy rate- 81% |
| code generation | McGowan M. et al. | Google Bard | Evaluate whether a code generated by Google Bard can improve the efficiency of quality assurance audits in gynaecological oncology | manual audits | Ovarian Cancer Audit- 600 surgical cases<br>Subspecialty Reaccreditation Audit - 390 surgical cases | Bard's ovarian cancer audit took less time per case compared to the manual audit, despite a larger number of cases. |
| medical education | Hermann CE. et al. | GPT-3.5 | Quantify the accuracy of GPT-3.5 in answering common questions pertaining to cervix cancer prevention | 2 Attending Gynecologic Oncologist | 64 questions | Correct and Comprehensive answers- 53%<br>Correct but Non-Comprehensive answers- 30%<br>Partially incorrect answers- 16%<br>Completely Incorrect answers- 2% |
| medical education | Patel JM. et al. | GPT-3.5 | Quantify the accuracy of GPT-3.5 in answering commonly asked questions pertaining to genetic testing and counseling for gynecologic cancers. | Two attending Gynecologic Oncologists | 40 questions | Correct and comprehensive answers- 82.5%<br>Correct and not comprehensive answers- 15%<br>Partially incorrect answers- 2.5%<br>Incorrect answers- 0% |

All studies were evaluated for risk of bias and applicability using the QUADAS-2 tool **(Supplementary Table 1)**. In medical education, one study^15^ showed a high risk in patient selection, while others generally exhibit low risk across criteria. Clinical practice showed a mix, with some studies^16^ having high risk in patient selection. In medical code generation, risks are mostly low except for some unclear assessments in the index test.

### Descriptive summary of results

#### Medical Education

Two studies assessed the efficacy of ChatGPT (GPT-3.5) in answering medical queries. Patel JM et al.^17^ evaluated the accuracy of GPT-3.5 in responding to commonly asked questions about genetic testing and counseling for gynecologic cancers. ChatGPT responses were evaluated by attending gynecologic oncologists for correctness and comprehensiveness, revealing that over 97% of ChatGPT’s answers were completely correct, 2.5% were partially correct, and none were completely incorrect. Conversely, Hermann CE et al.^15^ focused on cervical cancer prevention questions. The answers were similarly scored for accuracy and thoroughness by attending gynecologic oncologists. They found that only 83% of ChatGPT’s answers were completely correct, 16% were partially correct, and 2% were completely incorrect.

#### Clinical practice and therapeutic recommendation

Four studies evaluated the effectiveness of LLMs in clinical practice and therapeutic recommendations. Braun EM et al.^16^ assessed the ability of GPT-3.5 to recommend treatment for gynecological symptoms in palliative care scenarios. ChatGPT answers were evaluated by experts in gynecologic oncology and palliative care. The experts rated the guideline conformity of these recommendations with an average score of 4.1 out of 5. However, they noted that ChatGPT sometimes overlooked relevant therapies and failed to provide individualized assessments.

Meyer R et al.^18^ assessed the quality of postoperative instructions for gynecological procedures, comparing outputs from GPT-3.5, Google Search, and their institution’s standard instructions. The study revealed that GPT-3.5’s instructions had an understandability rate of 92%, comparable to both Google Search and the institution’s materials. However, the actionability rate for ChatGPT’s instructions was significantly lower at 60%, compared to the institution’s instructions.

Piazza. D. et al.^19^ conducted a comparative study examining the consistency and quality of responses generated by GPT-3.5 and GPT-4 in response to clinical queries about ovarian cancer, benchmarking them against the Italian Association of Medical Oncology (AIOM) guidelines. An expert panel of healthcare professionals evaluated the responses for clarity, consistency, comprehensiveness, usability, and overall quality using a five-point Likert scale. The study found that the AIOM guidelines significantly outperformed both GPT-3.5 and GPT-4 models, with no notable differences between the two GPT versions.

Krückel A. et al.^20^ evaluated ChatGPT’s ability to offer oncological treatment recommendations tailored to real, individual cases of endometrial, cervical, and ovarian cancers. Communications with ChatGPT were conducted in German, with scores ranging from -1 to 2. ChatGPT demonstrated promising results, with average scores of 0.75 for ovarian cancer, 0.7 for cervical, and 1.5 for endometrial cancer.

#### Medical Code Generation

Two studies utilized LLMs to generate code to determine if it could help streamline processes. The first study evaluated whether a code generated by Google Bard could improve the efficiency of quality assurance audits in gynecological oncology. It found that Bard’s ovarian cancer audit took less time compared to the manual audit.^21^ The second study investigated whether natural language processing (NLP) assisted by RoBERTa (based on Google’s BERT model) of unstructured operative notes could improve the prediction of residual disease in women with advanced epithelial ovarian cancer following cytoreductive surgery. The RoBERTa model outperformed models that used discrete clinical and engineered features and surpassed the performance of other state-of-the-art NLP tools.^22^

## DISCUSSION

LLMs are increasingly being researched in gynecologic oncology, covering areas such as medical education, clinical practice and code generation. However, they face challenges such as when tasked with understanding complex, debated medical practices. The studies reviewed demonstrate a range of strengths and weaknesses of LLMs in the field of gynecologic oncology, as detailed in **Table 3**. These issues emphasize the need for ongoing development.

**Table 3.**
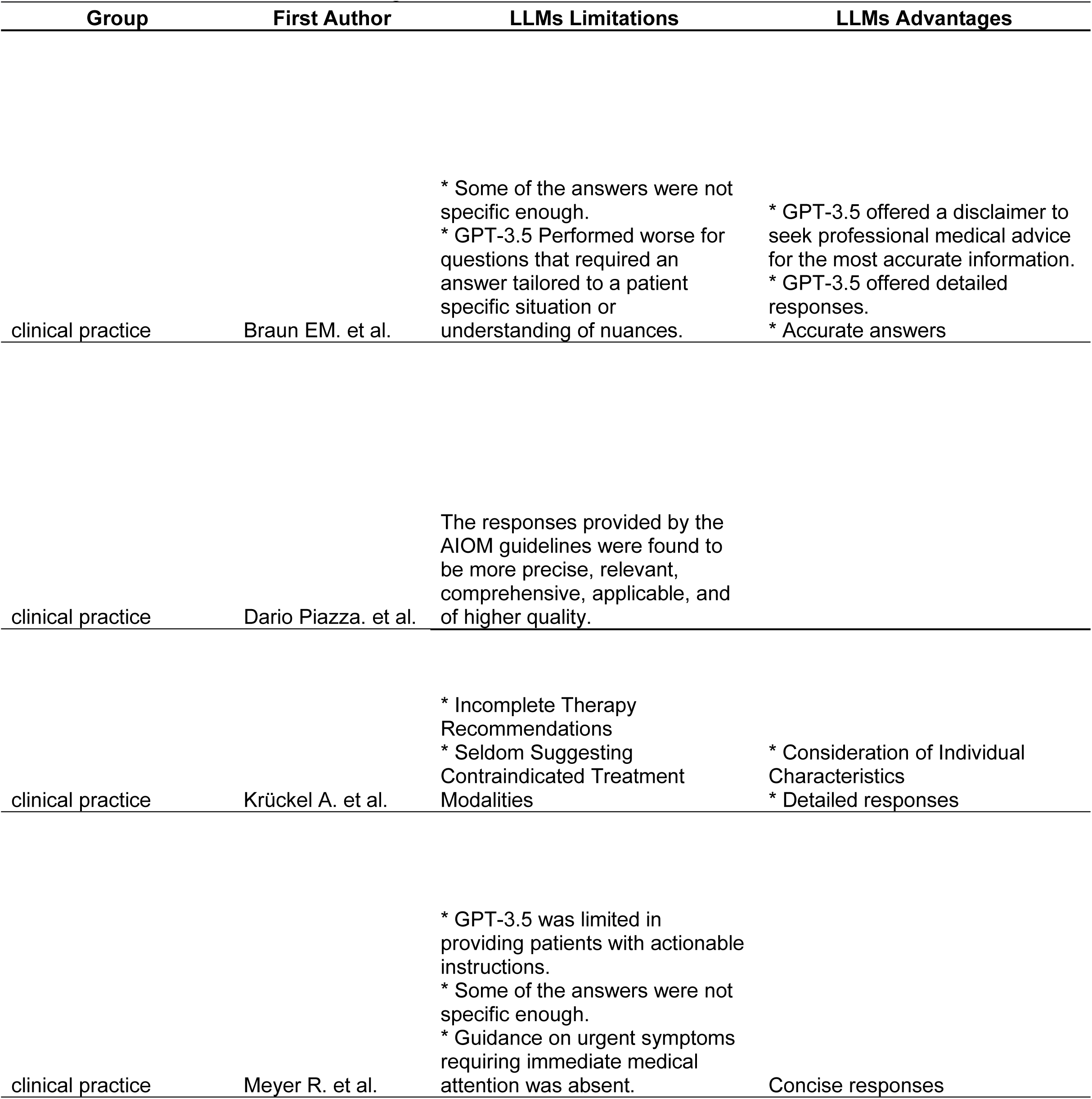
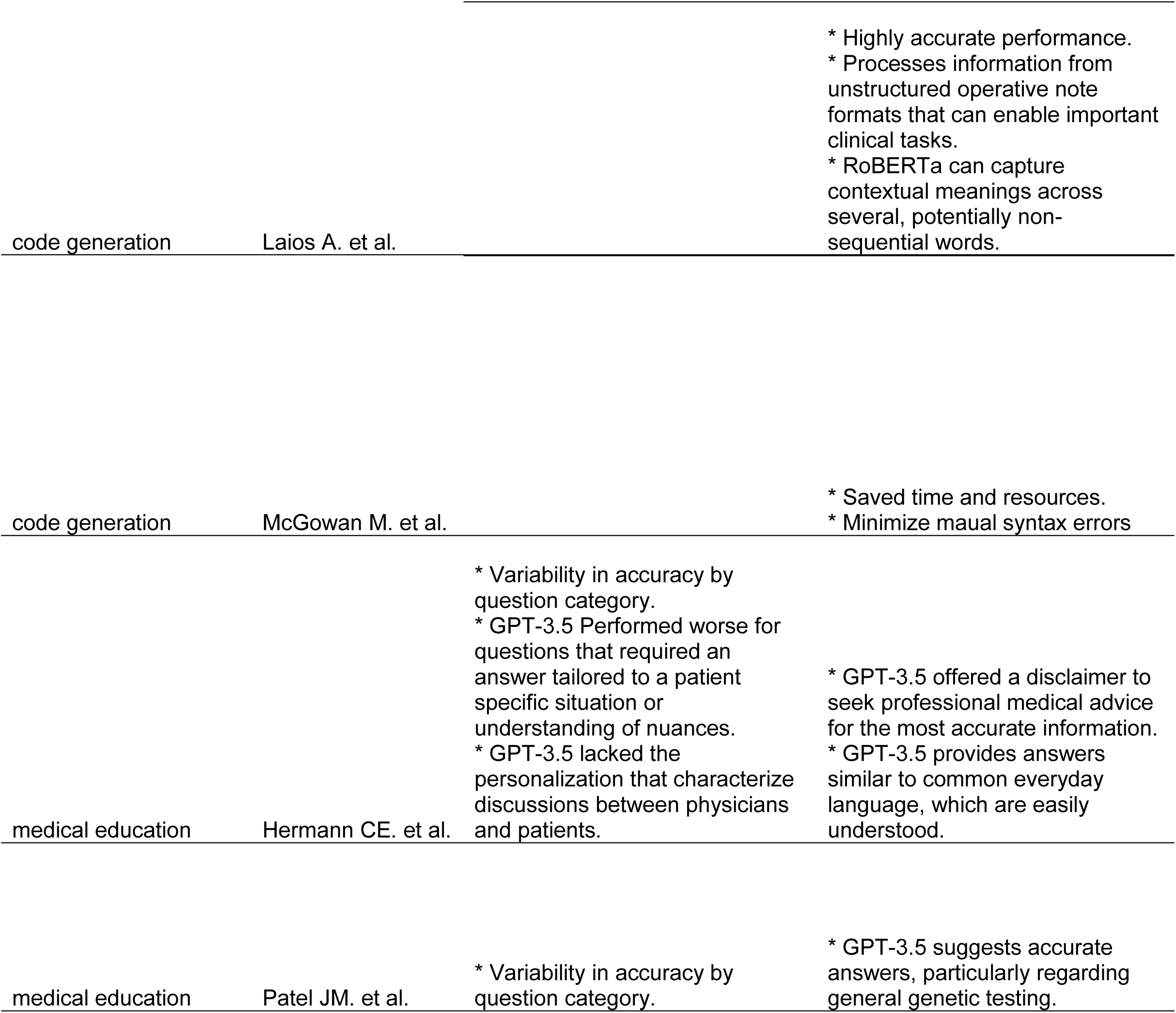
Limitations and Advantages of LLMs in the reviewed articles.

In medical education LLMs showed potential by providing accurate information.^17^ ^15^ For example, Patel JM et al.^17^ revealed ChatGPT’s efficacy in answering genetic testing and counseling queries, with a high accuracy rate. This suggests that LLMs can be reliable sources for fact-based medical education. However, Hermann CE et al.^15^ identified variability in performance, particularly noting a lower accuracy rate in cervical cancer prevention queries.

In providing medical recommendations, LLMs have demonstrated potential, yet they also exhibited significant limitations, mainly when tasked to comprhened individual patient characteristics .^16^ ^18^ ^19^ ^20^ Both Braun EM et al.^16^ and Krückel A. et al.^20^ observed that GPT-3.5 generally provided acceptable recommendations for palliative care and gynecological malignancies. However, Braun EM et al.^16^ also noted that the model’s recommendations often lacked personalized assessments. Furthermore, Meyer R et al.^18^ found that while GPT-3.5 generated understandable postoperative instructions, their practical applicability was limited. Additionally, guidelines from AIOM significantly surpassed both GPT-3.5 and GPT-4.^19^

The use of LLMs in medical code generation showcases their ability to streamline complex processes. The reviewed studies indicate that LLMs can surpass traditional electronic methods in medical coding.^21^ ^22^ For instance, Google Bard’s code significantly reduced the time needed for ovarian cancer quality assurance audits, saving both time and resources.^21^ Similarly, RoBERTa’s NLP capabilities outperformed traditional models in predicting residual disease post-surgery.^22^ These examples illustrate how LLMs can enhance operational efficiency and decision-making in gynecologic oncology by handling large datasets and performing intricate analyses. Also, these tools can make medical records more accessible to researchers, enabling them to perform higher quality studies.

LLMs show potential in gynecologic oncology, offering possibilities for both clinical practice and research. The models excel in analyzing large datasets, providing insights that can enhance patient care.^23^ ^24^ LLMs can assimilate published research and patient data to suggest up-to-date personalized treatment plans.^25^ Furthermore, LLMs can automate administrative tasks, such as creation of medical notes.^26^ ^27^ LLMs can also aid in medical research by identifying hidden patterns, potentially leading to new discoveries.^28^

Despite the promising applications of LLMs in gynecologic oncology, several significant challenges and limitations persist. A primary concern is data privacy, especially given the sensitive nature of gynecological oncological patient data.^29^ Another limitation is the interpretability of LLM outputs, as it can be challenging to understand the reasoning behind the model’s recommendations, which is important for clinical acceptanc.^30^ Furthermore, integrating these models into existing healthcare systems poses logistical challenges, including the need for continuous updates and maintenance.^31^ LLMs also risk providing responses that seem reasonable but are factually incorrect or irrelevant, known as ’hallucinations.’^32^

An impotnat aspect that remains overlooked by current litreture is the use of LLMs during the time of initial workup of suspected gynecologic malignancies prior to clear diagnosis. During this period LLM’s may serve to support paitents lacking immediate access to expert consultation.

Future research should prioritize refining LLM capabilities to address specific clinical challenges. Other subareas, such as uterine sarcomas, vaginal and vulvar cancer, have yet to be studied using LLMs. Even in more examined areas like cervical and ovarian cancers, existing research is still limited to a few studies. Systematic investigations across these various subareas will help to fully understand the capabilities and benefits of LLMs in gynecologic oncology. Studies aimed at enhancing the interpretability of these models are also important, as these qualities are needed for gaining trust among healthcare providers.

Out of the eight reviewed articles, six examined the performance of ChatGPT-3.5, one assessed the performance of Google Bard, one evaluated BERT, and one explored ChatGPT-4. (**Figure 4**) This demonstrate the necessity of examining different types of LLMs and comparing their performance in the field of gynecologic oncology.

**Figure 4.**
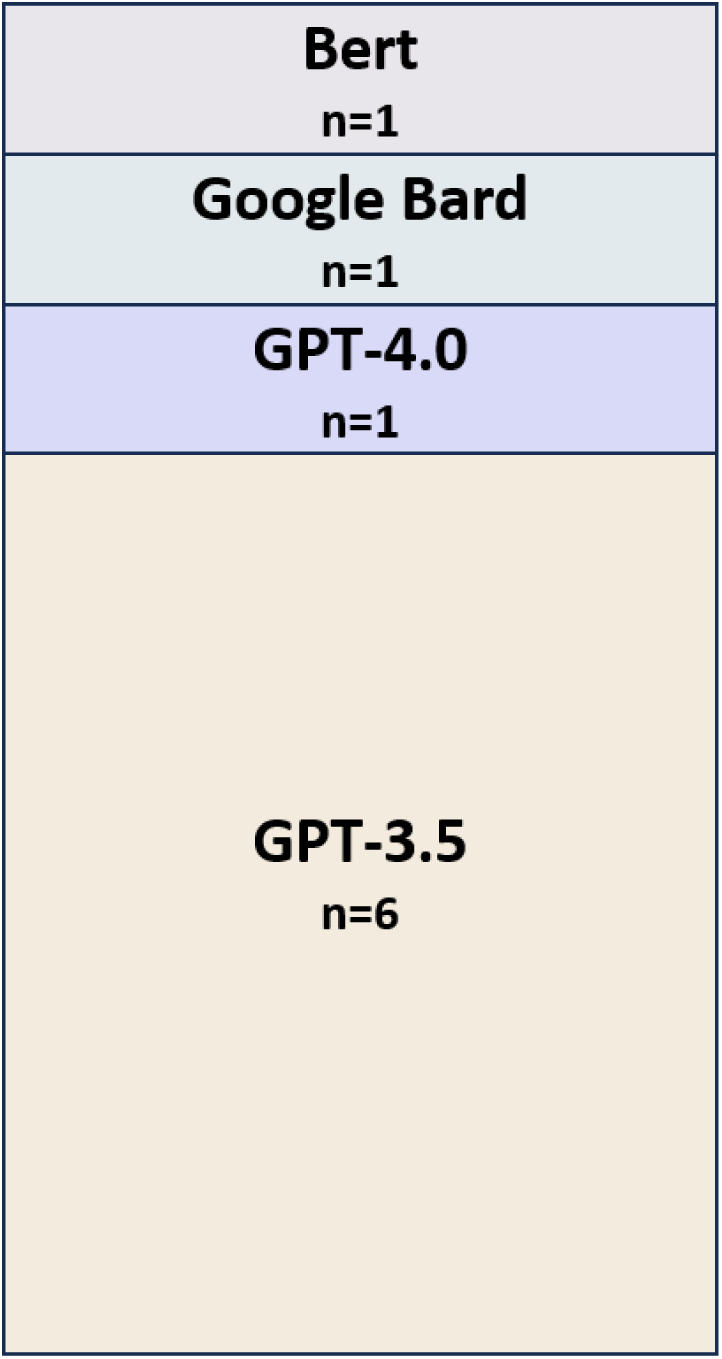
Number of reviewed articles according to the type of LLM used.

Our study faces several limitations. The inclusion of only eight studies does not provide a full view of the capabilities and limitations of LLMs in this area. Furthermore, the high risk of bias observed in some of the studies could affect the reliability of our findings. Moreover, the studies focus primarily on narrow aspects of gynecologic oncology, like genetic testing and palliative care, which may not be representative of other, more complex scenarios in the field. Another significant issue is the variability across the studies regarding their design, objectives, methodologies, and LLMs types. This diversity prevents forming consistent conclusions about the efficacy of LLMs across various applications within the specialty. Additionally, the rapid technological advancements in LLMs indicate that earlier studies might not reflect the current state of the technology. ^33^ ^34^ ^35^

In Conclusion, LLMs demonstrate inconsistent performance in gynecologic oncology, displaying both strengths and notable limitations. These findings emphasize the need for continuous evaluation of these models before they are implemented clinically.

## Data Availability

All data produced in the present work are contained in the manuscript

