## Supplementary for "Leveraging Large Language Models in Gynecologic Oncology: A Systematic Review of Current Applications and Challenges"

### Detailed Search Strategies

#### PubMed (PubMed.gov):

**#1**
(Gynecologic oncology) OR (Cervical cancer) OR (Endometrial cancer) OR (Ovarian cancer) OR (Fallopian tube cancer) OR (Vulvar cancer) OR (Vaginal cancer) OR (Uterine sarcoma) OR (Gestational trophoblastic disease) OR (HPV) OR (Cervical Cancer) OR (Gynecologic cancers) OR (Colposcopy) OR (Pap Smear)

**#2**
("ChatGPT") OR ("large language models") OR ("OpenAI") OR ("Microsoft bing") OR ("google bard") OR ("google gemini")

**#3**
(#1 AND #2) AND English **Filters:** from 2023 – 2024

#### Scopus (Scopus.com):

**#1**
(Gynecologic oncology) OR (Cervical cancer) OR (Endometrial cancer) OR (Ovarian cancer) OR (Fallopian tube cancer) OR (Vulvar cancer) OR (Vaginal cancer) OR (Uterine sarcoma) OR (Gestational trophoblastic disease) OR (HPV) OR (Cervical Cancer) OR (Gynecologic cancers) OR (Colposcopy) OR (Pap Smear)

**2#**
( "ChatGPT" ) OR ( "large language models" ) OR ( "OpenAI" ) OR ( "Microsoft bing" ) OR ( "google bard" ) OR ( "google gemini")

**#3**
(#1 AND #2) AND PUBYEAR > 2022 AND ( LIMIT-TO ( DOCTYPE , "ar" ) ) AND ( LIMIT-TO ( SUBJAREA , "MEDI" ) ) AND ( LIMIT-TO ( LANGUAGE , "English" ) )

#### Web of Science (Thomson Reuters):

**#1**
ALL=( Gynecologic oncology)) OR ALL=( Cervical cancer)) OR ALL=( Endometrial cancer)) OR ALL=( Ovarian cancer)) OR ALL=( Fallopian tube cancer)) OR ALL=( Vulvar cancer)) OR ALL=( Vaginal cancer)) OR ALL=( Uterine sarcoma)) OR ALL=( Gestational trophoblastic disease)) OR ALL=( HPV)) OR ALL=( Cervical Cancer)) OR ALL=( Gynecologic cancers)) OR ALL=( Colposcopy)) OR ALL=( Pap Smear))

**2#**
ALL=(ChatGPT)) OR ALL=(large language models)) OR ALL=(OpenAI)) OR ALL=(Microsoft bing)) OR ALL=(google bard)) OR ALL=(google gemini)

**#3**
(#1 AND #2)

#### Supplementary Figure 1- PRISMA flow diagram

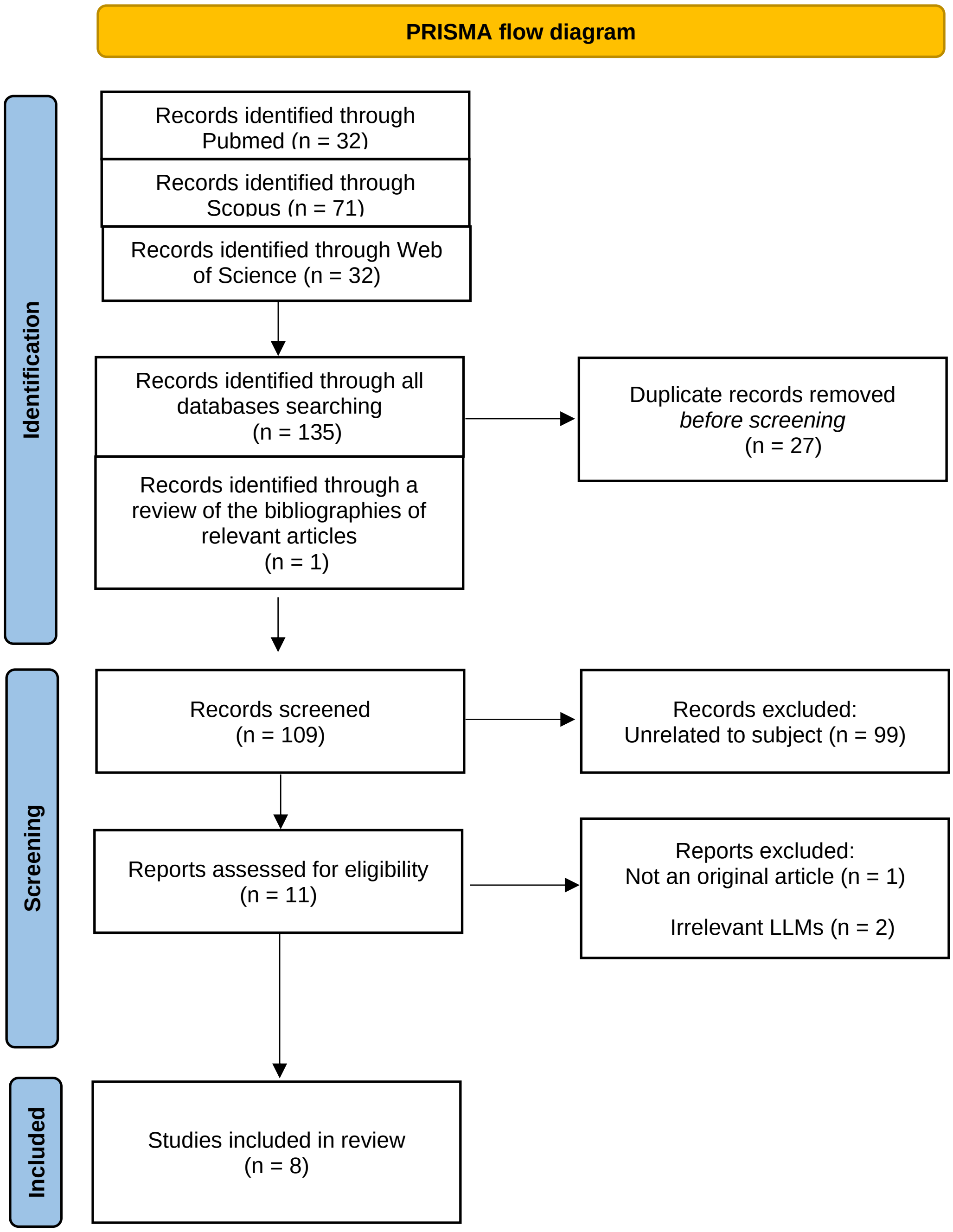


**Supplementary Figure 1.** Flow diagram of the search and inclusion process.

#### Supplementary Table 1- QUADAS-2 risk of bias

| Group | First Author | Patient Selection | Index Test | Reference Standard | Flow and timing |
|---|---|---|---|---|---|
| Medical Education | Hermann CE | High | Low | Low | Low |
| | Patel JM | Low | Low | Low | Low |
| Clinical practice | Braun EM | High | Low | Low | Low |
| | Piazza D | Low | Unclear | Low | Unclear |
| | Krückel A | Unclear | Low | Low | Unclear |
| | Meyer R | Unclear | Low | Unclear | Unclear |
| Medical Code Generation | Laios A | Low | Low | Low | Unclear |
| | McGowan M | Low | Unclear | Low | Low |

Low Risk of Bias    High Risk of Bias    Unclear Risk of Bias

**Supplementary Table 1.** QUADAS-2 risk of bias assessment per clinical application
